# A Descriptive Analysis of Chairs of Academic Neurology Departments in the United States

**DOI:** 10.1101/2021.11.06.21266013

**Authors:** Kajol K. Patel, Parth A. Patel

## Abstract

There remains a limited understanding of the characteristics of academic leaders within neurology departments, despite similar research in other fields. This investigation characterized the demographics, academic background, and scholarly productivity of United States (U.S.) neurology department chairs. Here, 131 chairs at Accreditation Council for Graduate Medical Education (ACGME)-certified neurology programs were identified. Publicly accessible demographic and academic data available online were collected in March 2021. Among the 131 neurology chairs analyzed, 84.7% were male. On average, these faculty were 60.5 years old and were appointed at a mean age of 52.0 years. 74.8% of chairs graduated from an American medical school, although a notable proportion of department heads received medical training internationally. A substantial cohort also acquired an additional graduate degree, of which Doctor of Philosophy (PhD; 22.1%) and master’s degree (21.4%) were most common. 82.4% completed a post-residency fellowship, which were most frequently in vascular neurology (24.1%) and clinical neurophysiology (17.6%). The mean *h-index, m-quotient*, and lifetime NIH grant funding received were 39.2 ± 29.4, 1.2 ± 0.8, and $20,021,594 ± $31,861,816, respectively. No between-gender differences were observed. Overall, neurology chairs are predominantly male, most often completing fellowships in vascular neurology or clinical neurophysiology. Research productivity is a notable component of these chairs’ careers, although certain programs place less emphasis on these metrics. Finally, substantial effort remains to address disparities in female representation at this leadership position. These findings serve as a benchmark to evaluate demographic trends among neurology department chairs.

## Introduction

Neurology department chairs represent important academic leaders within their respective institutions. Considering the substantial number of full-time neurology faculty at academic programs (U.S. Medical School Faculty by Department, 2020 2020), these individuals maintain significant responsibility as they are foundational for their respective department’s future accolades. However, despite this position’s importance, no research has investigated the characteristics of current neurology chairs in the United States (U.S.), including background and demographics. Such an analysis would facilitate a more representative understanding of the profiles of current leaders of academic neurology departments in the U.S. Therefore, the present study examines the academic background, demographic characteristics, and scholarly productivity of current chairs of U.S. academic neurology departments.

## Methods

A listing of academic neurology programs was obtained using the Fellowship and Residency Interactive Database Access System (FREIDA) website sponsored by the American Medical Association (AMA). Overall, 162 programs affiliated with the Accreditation Council for Graduate Medical Education (ACGME) were identified following exclusion of military programs. Each program’s chair was determined as the faculty member with any of the following titles: “Chair”, “Chairperson”, “Chief”, or “Head of Department”. Of note, 31 department heads could not be ascertained and therefore these programs were excluded from further analyses.

### Demographic Data Collection

For the remaining 131 programs, each chair’s information was acquired using publically available resources including departmental websites, online *curriculum vitae*, and program newsletters. Data were collected in March 2021 and included age, gender, medical school attended, graduation year, additional degrees earned, residency location, fellowship acquisition, year of appointment as chairperson, and status of the position (e.g. interim or permanent). The year of initial appointment could not be identified for seven chairpersons, who were subsequently excluded for those specific analyses. Information regarding age was collated using Healthgrades (https://www.healthgrades.com/). For three department heads, age was unavailable and therefore was estimated by adding 26 years (the mean age of medical school graduation in the U.S.) to each of their medical school graduation years, a methodology employed previously in the literature (Dotan et al. 2018).

Historically, fellowship programs have been altered and structural changes have been implemented. Therefore, for the present analyses, certain subspecialties were categorized together to maintain uniformity. Cerebrovascular disease, stroke, and vascular neurology were collectively defined as vascular neurology. Electromyogram (EMG), electroencephalogram (EEG), epilepsy, and clinical neurophysiology were collectively defined as clinical neurophysiology.

### Scholarly Productivity

Scholarly activity was evaluated through the Hirsch index (*h-index*), the *m-quotient*, and total National Institutes of Health (NIH) grant funding received as principal investigator. The *h-index* is defined as the largest number of papers, *h*, for which the citation count exceeds or is equal to *h* (Hirsch 2005). For example, if a researcher has 15 publications, each with at least 15 citations, then their *h-index* is defined as 15. The *m-quotient* is a modification that is derived from dividing the *h-index* by the number of years since initial publication, thereby accounting for variation in career duration. Although multiple methods exist for acquiring the *h-index*, the present study utilized Scopus (https://www.scopus.com), which has previously been demonstrated to correlate with other databases such as Google Scholar and Web of Science (Harzing and Alakangas 2016). Each chair’s year of first publication was tabulated by entering their first initial, middle initial (where available), and last name into the National Library of Medicine PubMed website (https://www.ncbi.nlm.nih.gov/pubmed).

The NIH Research Portfolio Online Reporting Tools Expenditures and Reports (RePORTER) database (https://reporter.nih.gov/) was queried to obtain total lifetime grant funding amounts for each department head. This tool provides NIH funding information between 1985 and the present, with all years of available data included for the present study.

### Ethical Considerations

This study did not collect protected patient health information and therefore Institutional Review Board approval was not required.

### Statistical Analysis

As data were normally distributed on visual inspection, Student’s t-tests were used for between-group comparisons with a threshold for significance set at *P <* 0.05. All descriptive and statistical analysis was performed using GraphPad Prism 8.4.2 (San Diego, CA).

## Results

### Demographic Characteristics

Among the 131 neurology departments analyzed, it was observed that 111 (84.7%) chairs were male and 20 (15.3%) were female. Currently, the average age of department heads is 60.5 (standard deviation [s.d.]) ± 8.1 years (median, 60 years; interquartile range [IQR], 55 – 66 years). The majority of chairs are between 60 and 69 years old (42.0%) and 50 and 59 years old (38.2%) (Figure 1). These chairs were appointed to the position at a mean age of 52.0 (s.d.) ± 6.7 years (median, 52 years; IQR, 48 – 57 years) following an average of 26.2 (s.d.) ± 6.9 years (median, 26; IQR, 22 – 42) post-medical school completion. Each current chairperson has served 8.5 (s.d.) ± 7.3 years (median, 7 years; IQR, 3 – 12 years), on average, and 50 (40.3%) were appointed within the past five years (Figure 2). 124 chairs (94.7%) currently occupy a permanent position, whereas 7 chairs (5.3%) are currently of interim status.

**Figure 1:**
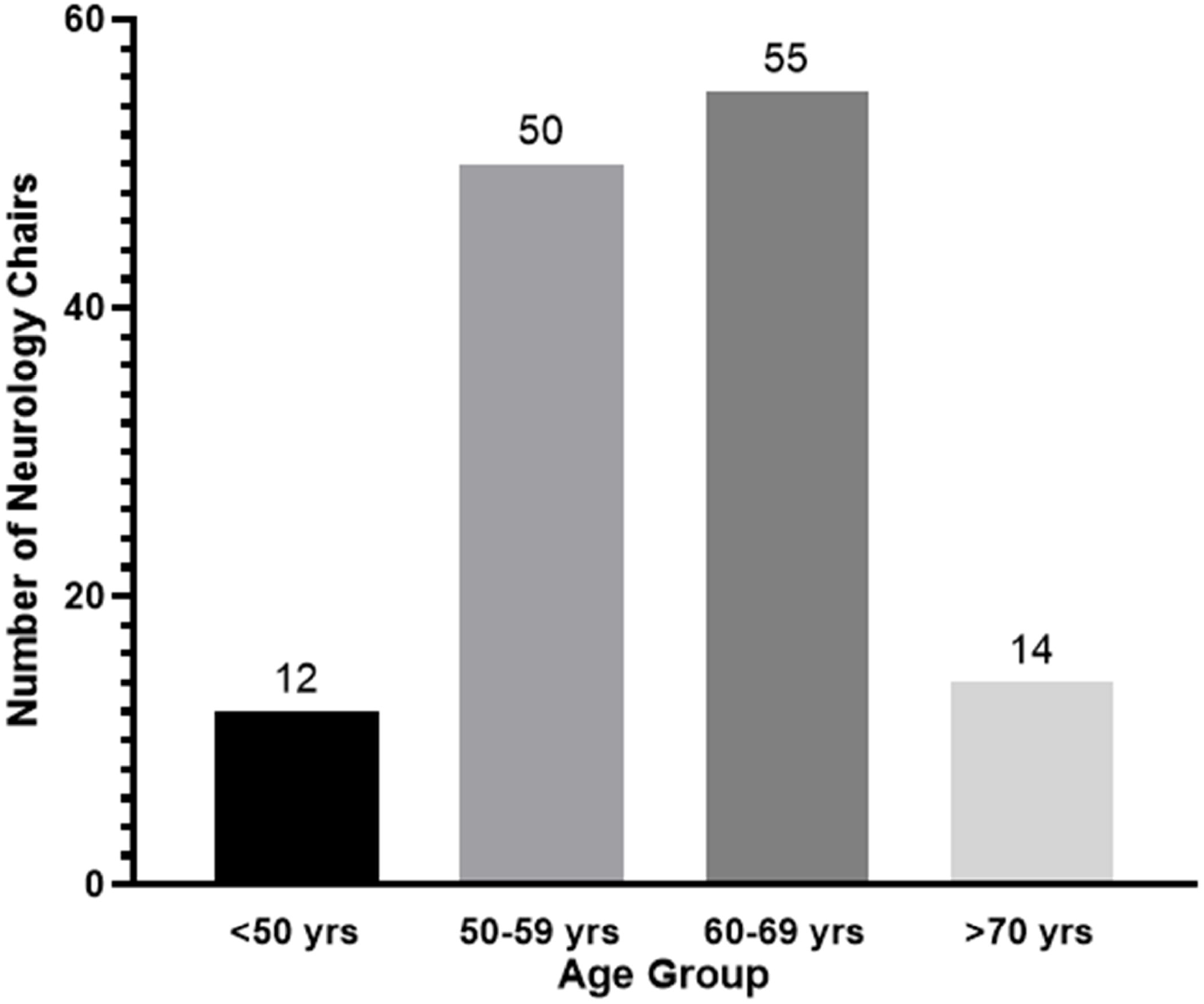
Distribution of current neurology chairs by age.

**Figure 2:**
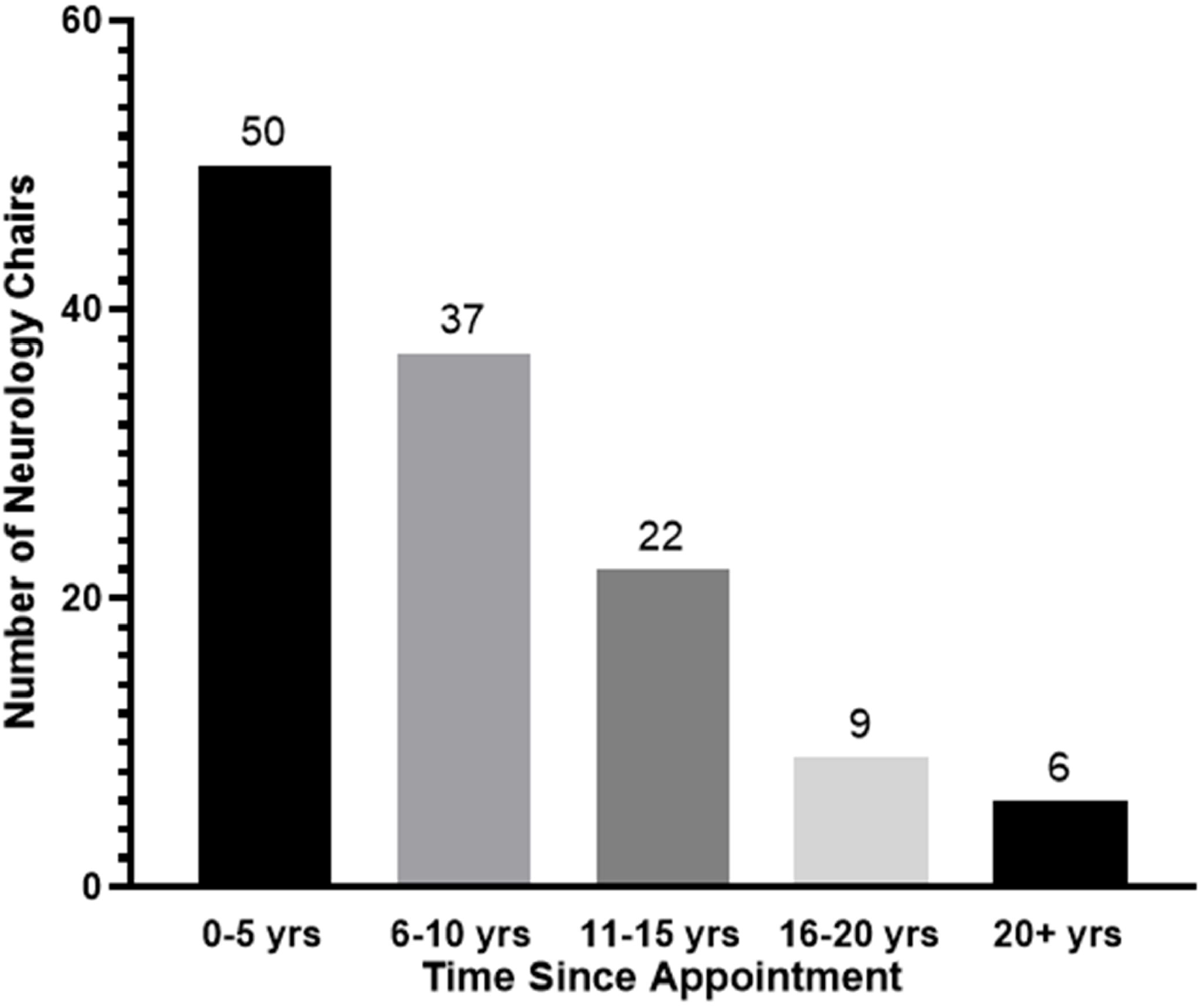
Distribution of current neurology chairs by years since initial appointment.

### Academic Background

Although the majority of department heads completed medical school in the U.S. (98 of 131 [74.8%]), a substantial number graduated from an international medical program (33 of 131 [25.2%]). The locations of these international medical schools were myriad, but the largest proportion of these faculty attended a program in India (6 of 33 [18.2%]). Among American medical schools, graduates from Washington University School of Medicine in St. Louis and Johns Hopkins University School of Medicine comprised the greatest proportion of department heads (7 of 131 [5.3%] and 6 of 131 [4.6%], respectively; Table 1). Most department chairs (128 of 131 [97.7%]) completed a Medical Doctor (MD) degree, with the remaining chairs (3 of 131 [3.3%]) completing a Doctor of Osteopathy (DO) degree. 39.7% (52 of 131) of chairs possess additional graduate degrees, the most common of which are a Doctor of Philosophy (PhD; 29 of 131 [22.1%]) and master’s degree (28 of 131 [21.4%]).

**Table 1:** Medical Schools, Residency Programs, and Fellowship Programs Attended by 3 or More Current Chairs of Neurology Departments

| Medical School (State) | Chairs |
| --- | --- |
| Washington University School of Medicine (MO) | 7 |
| Johns Hopkins University (MD) | 6 |
| Albert Einstein (NY) | 3 |
| Boston University (MA) | 3 |
| Cornell University (NY) | 3 |
| Northwestern (IL) | 3 |
| Stanford (CA) | 3 |
| West Virginia University (WV) | 3 |
| Residency Program (State) | Chairs |
| Massachusetts General Brigham (MA) | 8 |
| Johns Hopkins University (MD) | 7 |
| Columbia University (NY) | 6 |
| Mount Sinai (NY) | 6 |
| Mayo Clinic (MI) | 5 |
| University of Pennsylvania (PA) | 5 |
| Washington University School of Medicine (MO) | 5 |
| Cornell University (NY) | 4 |
| University of California San Francisco (CA) | 4 |
| Albert Einstein (NY) | 3 |
| University of Chicago (IL) | 3 |
| Fellowship Program (State) | Chairs |
| Johns Hopkins University (MD) | 8 |
| Massachusetts General Hospital (MA) | 7 |
| Duke University (NC) | 5 |
| Columbia University (NY) | 4 |
| University of California San Francisco (CA) | 4 |
| Baylor University (TX) | 3 |
| Brigham and Women's Hospital (MA) | 3 |
| Cleveland Clinic (OH) | 3 |
| Harvard University (MA) | 3 |
| Mayo Clinic (MI) | 3 |
| National Institutes of Health (MD) | 3 |
| University of California Los Angeles (CA) | 3 |

With the exception of 1 chairperson that exclusively completed residency training internationally, all department heads completed neurology residency training in the United States. The most common programs among this cohort included the combined Massachusetts General Brigham neurology residency (8 of 131 [6.1%]; formerly Massachusetts General Hospital and Brigham and Women’s Hospital) and Johns Hopkins University neurology residency (7 of 131 [6.1%]) (Table 1). A further 108 chairs (82.4%) completed additional post-residency fellowship training, of whom 20 (15.3%) completed two or more distinct fellowships. These were most frequently acquired at Johns Hopkins University (8 of 108 [7.4%]) and Massachusetts General Hospital (7 of 108 [6.5%]) (Table 1). Clinical and/or research fellowship programs were most commonly completed in vascular neurology (26 of 108 [24.1%]) and clinical neurophysiology (24 of 108 [22.2%]) (Figure 3).

**Figure 3:**
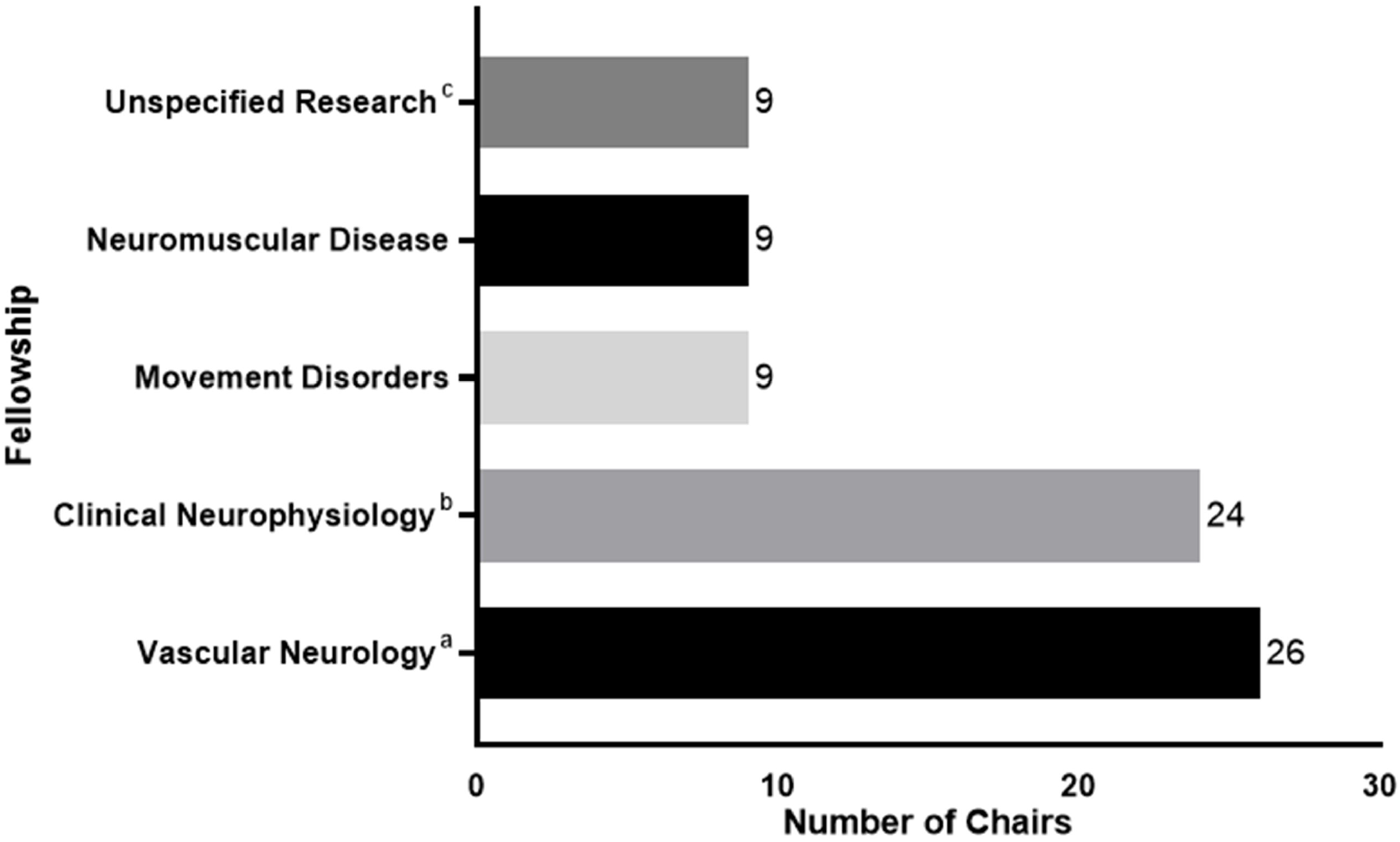
Top five most common fellowships for current neurology chairs. ^a^Vascular neurology includes clinical and/or research fellowships in cerebrovascular disease, stroke, and vascular neurology. ^b^Clinical neurophysiology includes electromyogram (EMG), electroencephalogram (EEG), epilepsy, and clinical neurophysiology. ^c^Unspecified research is defined as research fellowships that were not categorized e.g., as vascular neurology, clinical neurophysiology, etc.

### Scholarly Productivity

An assessment of the scholarly productivity of neurology chairs indicated a mean *h-index* and *m-quotient* of 39.2 (s.d.) ± 29.4 (median, 31.0; IQR, 19.0 – 55.0) and 1.2 (s.d.) ± 0.8 (median, 1.1; IQR 0.6 – 1.6), respectively. The majority of these faculty (73 of 131 [55.7%]) received NIH grant funding during their careers; of these department heads, the average total lifetime amount of funding received was $20,021,594 (s.d.) ± $31,861,816 (median, $9,328,086; IQR, $2,345,289 – $21,137,970). Following stratification by gender, no significant between-group differences were observed for *h-index* (males, 38.8 [s.d.] ± 30.3 versus females, 41.1 [s.d.] ± 24.4; *P* = 0.76) and *m-quotient* (males, 1.2 [s.d.] ± 0.9 versus females, 1.3 [s.d.] ± 0.8; *P* = 0.8). Furthermore, among NIH-funded faculty, males and females received similar amounts of total lifetime funding ($20,813,224 [s.d.] ± $34,007,721 versus $16,685,439 [s.d.] ± $21,161,239, respectively; *P* = 0.67).

## Discussion

The present study provided an evaluation of the demographic characteristics, academic backgrounds, and scholarly productivity of 131 neurology department chairs. Such a characterization should provide a useful benchmark for understanding the current state of leadership in academic neurology and for temporal comparisons of changes in the composition of these leaders.

Expectedly, males comprised the majority of neurology chairs (85%). Neurology has historically been dominated by males, with recent estimates suggesting that females constitute only 30.9% of active neurologists (Active Physicians by Sex and Specialty, 2019 2019). This gender gap extends to academic medicine, with a disparity that increases with advanced academic rank. The findings described here are consistent with the latter results, which demonstrated that only 13.8% of neurology professors were female (McDermott et al. 2018). As such, males possess departmental leadership positions at a disproportionately greater frequency to their overall representation in the field. These observations are troubling considering female neurologists already report substantially reduced compensation (Zecavati et al. 2016), decreased academic output (McDermott et al. 2018), and less representation among leadership of professional societies (Silver et al. 2019). Despite these biases, the gender composition of the field is evolving, as women constituted 45.9% of incoming neurology residents in 2020 (Table B3. Number of Active Residents, by Type of Medical School, GME Specialty, and Sex 2020). This transition is encouraging, and as more females occupy senior faculty positions, we expect the proportion of female neurology chairs to correspondingly increase.

Neurology chairs were initially appointed to their respective positions at a mean and median age of 52 years, which is comparable to the findings of previous studies in specialties such as general surgery (49 years) (Shuck 2002), ophthalmology (47 years) (Dotan et al. 2018), and urology (47 years) (Farber et al. 2017). Considering the immense responsibilities associated with this position, the advanced average age of appointment to this position coheres with the expectation for these faculty members to be well-established professionally.

In addition to their medical degree, 39.7% of chairs possess an advanced academic degree. The career stage during which these degrees were completed (i.e., during and/or around completion of medical degree, immediately prior to appointment as chair, or following accession to department head) was unable to be distinguished. Nonetheless, compared to both U.S. MD seniors who matched into neurology in 2020 (11.1%) (Charting Outcomes in the Match, 2020 2020), and academic leaders within specialties such as ophthalmology (8%) (Dotan et al. 2018) and urology (5%) (Farber et al. 2017), a substantially larger proportion of neurology chairs completed a PhD (22.1%). Therefore, relative to other fields, acquisition of this terminal degree may be an influential factor in appointment to this position.

Although the majority of department heads in this study graduated from a U.S. medical school (74.8%), a substantial minority completed their medical education internationally (25.2%). This is a notably greater proportion compared to ophthalmology, where only 8% of chairpersons graduated from an international medical school (Dotan et al. 2018). These findings may partially be attributed to differences in the compositions of both fields: in 2019, 31.5% of neurologists were international medical graduates (IMGs), compared to 6.8% of ophthalmologists (Active Physicians Who Are International Medical Graduates (IMGs) by Specialty, 2019 2019). Among individuals with medical training in the United States, the most frequently attended medical school was Johns Hopkins University. Interestingly, Johns Hopkins University was similarly a common site for residency and fellowship training, in addition to programs affiliated with Harvard University (Massachusetts General Hospital and Brigham and Women’s Hospital). Based on the present analysis, it is difficult to ascertain the reasons for specific programs to produce department leaders – that is, whether these institutions simply recruit more “well-qualified” candidates or whether they provide an environment that stimulates development of leadership ambitions and qualities.

Among neurology chairs with fellowship training, the most commonly completed programs were in vascular neurology and clinical neurophysiology (which further included EEG, EMG, and epilepsy). Considering a vascular neurology fellowship is offered by 106 programs, a clinical neurophysiology fellowship offered by 92 programs, and an epilepsy fellowship offered by 81 programs, as of 2020 (ACGME Data Resource Book 2021), it is not surprising that these are the most prevalent subspecialties.

Prior literature has established a robust relationship between the scholarly productivity of the chair and their academic department (Stavrakis et al. 2015; Zelle et al. 2017).

Therefore, for departments oriented towards maximizing research output, appointing a similarly research-focused clinician would cohere with this objective. Indeed, as demonstrated by the results of this study, scholarly productivity is an integral component of promotion to department chair. The mean and median *h-index* among this cohort were 39.2 and 31.0, respectively, which, on average, are greater than those reported for other specialties including anesthesiology (18) (Pagel and Hudetz 2011), ophthalmology (24) (Dotan et al. 2018), otolaryngology (16) (Eloy et al. 2015), and urology (31) (Farber et al. 2017). Irrespective of this overall finding, it is imperative to note that a large degree of variability was observed for this metric, suggesting that despite its significance for academic promotion, there exist substantial differences in programs’ perspectives towards research productivity. That is, certain programs may value criteria unrelated to scholarly output when evaluating candidates for the position of chair. Nonetheless, provided the findings described here, we speculate that research productivity is of considerable importance for appointment to neurology chair, perhaps to a greater extent than among other specialties.

The *m-quotient* and total NIH grant funding are additional proxies of academic output that were reported in the present study. Here, it was calculated that the mean and median *m-quotient* for neurology chairs was 1.2 and 1.1, respectively, which is greater than the median *m-quotients* for urology (0.97) (Mayer et al. 2017) and neurological surgery (1.0) (Khan et al. 2014). In addition, NIH-funded faculty received an average of $31,861,816 in total lifetime grant funding. These metrics further corroborate the substantial scholarly productivity of neurology department heads.

Somewhat surprisingly, stratification by gender revealed no significant differences with respect to measures of academic output. Previous literature in neurology has demonstrated that males produce more publications compared to their female counterparts throughout all academic ranks (McDermott et al. 2018). However, in that investigation, the authors did not discriminate between professor and chairpersons. The findings described here suggest that these gender differences do not extend to the latter position. Although the reasons for this phenomenon are beyond the scope of the present study, we hypothesize that males are initially more productive, but during the later stages of their careers, are surpassed by their female colleagues. This discrepancy may partially be explained by the disproportionate share of familial obligations assumed by female physicians, thereby potentially decreasing their initial productivity (Kuehn 2012; Reed and Buddeberg-Fischer 2001). Indeed, investigations in other specialties including ophthalmology (Dotan et al. 2018) and orthopedic surgery (Bastian et al. 2017) have demonstrated a similar absence of gender differences in the *h-index* among their respective department chairs. Nevertheless, despite parallel observations among other fields (Eloy et al. 2012; Lopez et al. 2014), this trend merits further investigation specifically with regards to academic neurology.

There are several limitations to note in this study. Information was collected using online resources and was not validated through contact with each included department. Indicative of this deficiency, the department heads for 31 programs were unable to be identified and the initial year of appointment was difficult to obtain for 7 chairs. However, provided the potential for response bias that may have occurred through the use of a prospective survey, the present analyses provide a more objective characterization of current neurology chairs. Furthermore, to determine the year of initial publication, each researcher was queried using PubMed, a database that may potentially provide results for multiple homonymous authors. To avoid this predicament, the middle initial, where available, was included for searches, similar to analyses in other fields (Dotan et al. 2018; Kloosterboer et al. 2020). If possible, this information was further confirmed through cross-references with publically accessible *curriculum vitae*. Finally, as a descriptive, rather than comparative, analysis, the intention of the present study was not to derive conclusions regarding differences between chairs and non-chairs. Further investigation would be needed for this purpose.

## Conclusion

Overall, the present study demonstrates that the majority of neurology chairs are males who graduated from U.S. allopathic institutions, although, compared to other specialties, there exists a substantial internationally-trained cohort. Interestingly, a considerable proportion of these academic leaders possess an additional degree, most commonly a PhD, thereby potentially suggesting the benefit of such supplementary training for accession to department head. Moreover, as assessed by the *h-index, m-quotient*, and total lifetime NIH grant funding, neurology chairs are prolific scholars. However, this does not appear to be the most important criterion for appointment at every institution. In general, while the findings of this investigation provide insight into the academic and professional backgrounds of current neurology chairs, future research should strive to further explore evolving trends at this leadership position. Such studies would provide additional guidance to academic neurologists aspiring to this role.

## Declarations

### Availability of data and material

All data are available upon request.

## Acknowledgements

None.

## Abbreviations

NIH: National Institutes of Health
AMA: American Medical Association
PhD: Doctor of Philosophy

